# Optimize Clinical Laboratory Diagnosis of COVID-19 from Suspect Cases by Likelihood Ratio of SARS-CoV-2 IgM and IgG antibody

**DOI:** 10.1101/2020.04.07.20053660

**Authors:** Feng Yangchun

## Abstract

**Objective:** To optimize clinical laboratory diagnosis of COVID-19 from suspect cases by Likelihood Ratio of SARS-CoV-2 IgM and IgG antibody.

**Methods:** By reinterpreting the data in the article “Diagnostic Value of Combined Detection of Serum 2019 novel coronavirus IgM and IgG Antibodies in novel coronavirusin Infection”, the positive likelihood ratio of IgM and IgG antibody in diagnosis of COVID-19 (nucleic acid positive patients) was calculated, and the posterior probability of IgM and IgG antibodies and their tandem detection to diagnose was finally calculated.

**Results:** The positive likelihood ratios of single IgM and IgG antibody were 18.50 and 12.65 respectively, and the posterior probabilities were 90.18% and 86.26% respectively. However, the posterior probability of the two antibodies tandem detection is 99.15%, which can give clinicians quantitative confidence in the diagnosis of COVID-19 from suspected cases. According to the results of this study, combining the advantages and disadvantages of nucleic acid detection and antibody detection, the clinical pathway for clinicians to diagnose COVID-19 is found.

**Conclusion:** For suspected cases, IgM and IgG antibody tests should be firstly done at the same time. If the antibody tests are all positive, COVID-19 can be confirmed. If not, nucleic acid detection (one or more times) is performed, and in extreme cases, high-throughput viral genome sequencing is performed.

---

Since the epidemic outbreak of Severe Acute Respiratory Syndrome Coronavirus 2 (SARS-CoV-2) from last December in China, more than one million cases had been confirmed in more than 150 countries including China. As the country with the most severe epidemic, China has accumulated a lot of experience in the diagnosis and treatment of COVID-19^[1-2]^. Based on increased awareness of the disease, 7th version of the new treatment plan for COVID-19 have been released by Chinese government. In the latest version^[3]^, which is “Diagnosis and Treatment Protocol for Novel Coronavirus Pneumonia (Trial Version 7)”, the standard indicators used for confirmed diagnosis of suspected cases including nucleic acid detection, gene sequencing and antibody detection of SARS-CoV-2. Among them, antibody detection was first written into the diagnostic criteria. On April 2nd, FDA also approved the first SARS-CoV-2 antibody test kit for COVID-19 detection. How to use nucleic acid detection and antibody detection to diagnose suspected COVID-19 patients more efficiently is a question worth pondering. So this study intends to use likelihood ratio to optimize the clinical pathway for clinicians to diagnose suspected cases by nucleic acid and antibody detection.

## Data and Methods

### 1. Information

The data of this study come from the articles about COVID-19 published online by Chinese Journal of Laboratory Medicine. It mainly includes the relevant data mentioned in “Analysis of novel coronavirus Nucleic Acid Detection and Co-infection Results of 8274 Subjects in Wuhan Area”(DOI: 10.3760/CMA.J.CN 114452- 2020 222-00106) ^[4]^and “Diagnostic Value of Combined Detection of novel coronavirus IgM and IgG Antibody in novel coronavirus Infection by Serum 2019 Method and Data Analysis”(DOI:10.3760/CMA.J.CN 114452-2020223-00109)^[5]^. The first author organization of the two articles are the Laboratory Medicine Center of People’s Hospital Affliated to Wuhan University. The subjects were all suspected and confirmed cases of COVID-19 admitted to the people’s hospital Affliated to Wuhan University.

### 2. Case Definitions

#### 2.1 Suspect cases

Considering both the following epidemiological history and clinical manifestations.

1. Epidemiological history: History of travel to or residence in Wuhan and its surrounding areas, or in other communities where cases have been reported within 14 days prior to the onset of the disease; In contact with novel coronavirus infected people (with positive results for the nucleic acid test) within 14 days prior to the onset of the disease;In contact with patients who have fever or respiratory symptoms from Wuhan and its surrounding area, or from communities where confirmed cases have been reported within 14 days before the onset of the disease; Clustered cases (2 or more cases with fever and/or respiratory symptoms in a small area such families, offices, schools etc within 2 weeks).
2. Clinical manifestations: Fever and/or respiratory symptoms; The aforementioned imaging characteristics of COVID-19; Normal or decreased WBC count, normal or decreased lymphocyte count in the early stage of onset. A suspect case has any of the epidemiological history plus any two clinical manifestations or all three clinical manifestations if there is no clear epidemiological history.

#### 2.2 Confirmed cases

Suspect cases with one of the following etiological or serological evidences: (1)Real-time fluorescent (RT-PCR) indicates positive for SARS-CoV-2 nucleic acid; (2)Viral gene sequence is highly homologous to known SARS-CoV-2. (3)SARS-CoV-2 specific IgM and IgG are detectable in serum; SARS-CoV-2 virus specific IgG is detectable or reaches a titration of at least 4-fold increase during convalescence compared with the acute phase.

### 3. Data and Test

The positive Likelihood Ratio (LR+) of IgM and IgG in suspected cases for nucleic acid positive patients was calculated mainly by reinterpreting the data in the article “Diagnostic Value of Combined Detection of Serum 2019 novel coronavirus IgM and IgG Antibody in novel coronavirus Infection”, According to the relevant data in “novel coronavirus Nucleic Acid Detection and Analysis of Combined Infection Results of 8274 Subjects in Wuhan Area”, the posterior probability of IgM and IgG antibody detection alone and in series for diagnosis of nucleic acid positive patients was calculated. The SARS-CoV-2 IgM and IgG are tested by chemiluminescence immune assay(CLIA), the nucleic acid also are tested by RT-PCR.

## Results

### 1. Diagnostic performance of IgG and IgM in diagnosing COVID-19

According to the article “Diagnostic Value of Combined Detection of Serum 2019 novel coronavirus IgM and IgG Antibodies in novel coronavirus Infection”, 186 patients with positive nucleic acid detection were mentioned, and 98 patients with negative nucleic acid detection. Combined with the number of IgG and IgM antibody detection cases mentioned in the article, Table 1-2 was obtained.

**Table 1:** Comparison of SARS-CoV-2 IgG Antibody and Nucleic Acid Test.

| SARS-CoV-2<br>IgG Test | SARS-CoV-2 Nucleic Acid Test |  | Total |
| --- | --- | --- | --- |
|  | Positive | Negative |  |
| Positive | 197 | 6 | 205 |
| Negative | 8 | 73 | 81 |
| Total | 205 | 79 | 284 |

**Table 2:** Comparison of SARS-CoV-2 IgM Antibody and Nucleic Acid Test.

| SARS-CoV-2<br>IgM Test | SARS-CoV-2 Nucleic Acid Test |  | Total |
| --- | --- | --- | --- |
|  | Positive | Negative |  |
| Positive | 144 | 3 | 147 |
| Negative | 61 | 76 | 137 |
| Total | 205 | 79 | 284 |

From Table 1, it can be calculated that the sensitivity, specificity, negative predictive value and positive predictive value of SARS-CoV-2 IgG antibody detection to diagnose COVID-19 are 96.10%, 92.40%, 90.10% and 96.09%, respectively. The negative likelihood ratio is 0.04 and the positive likelihood ratio is 12.65. The accuracy is 95.07%.

From Table 2, it can be calculated that the sensitivity, specificity, negative predictive value and positive predictive value of SARS-CoV-2 IgM antibody detection to diagnose COVID-19 are 70.24%, 96.20%, 56.72% and 97.76%, respectively. The negative likelihood ratio is 0.31 and the positive likelihood ratio is 18.50. The accuracy is 77.46%.

### 2. Posterior Probability of Single IgG, IgM and IgM/IgG tandem diagnostic test

The posterior probability formula of single index:

> Prior ratio = prior probability/(1–prior probability)
>
> Posterior ratio = Prior ratio ×LR+
>
> Posterior probability = posterior ratio /(1+ posterior ratio)

The posterior probability formula of tandem diagnostic test of multiple indexes:

> Prior ratio = prior probability/(1–prior probability)
>
> Posterior ratio = Prior ratio ×LR+ _1_×LR+_2_×…×LR+_n_
>
> Posterior probability = posterior ratio /(1+ posterior ratio)

According to the 8274 suspected cases of COVID-19 explicitly mentioned in “Analysis of novel coronavirus Nucleic Acid Detection and Co-infection Results of 8274 Subjects in Wuhan Area”, the proportion of confirmed cases from suspected cases is 33.17%, which is the prior probability. The posterior probability of single test of IgG, IgM and IgM/IgG tandem diagnostic test were on Table 3.

**Table 3:** Posterior Probability of Diagnostic COVID-19 by Single IgG, IgM Test and IgM/IgG Tandem Test.

|  | Prior Probability | Prior Ratio | Posterior Ratio | Posterior Probability |
| --- | --- | --- | --- | --- |
| IgG Test | 33.17% | 0.4963 | 6.28 | 86.26% |
| IgM Test | 33.17% | 0.4963 | 9.18 | 90.18% |
| IgM/IgG Tandem Test | 33.17% | 0.4963 | 116.19 | 99.15% |

For suspected cases, after single detection of IgG and IgM antibodies, the clinician’s confidence in the diagnosis of suspected patients as COVID-19 confirmed cases is between 86.26% and 90.18%, but after IgM/IgG tandem test, the clinician’s confidence in the diagnosis of suspected patients as COVID-19 confirmed cases has increased to 99.15%.

### 3. Clinical Pathway for Clinicians to Diagnose COVID-19 Based on IgM/IgG Antibody Tandem Test

According to the results of this study, combining the advantages and disadvantages of nucleic acid detection and antibody detection, the clinical pathway for clinicians to diagnose COVID-19 is shown in Figure 1. It proposed a clinical path for diagnostic COVID-19, starting with IgM/IgG tandem test, and with subsequent use, if needed, of nucleic acid test and then high-throughput sequencing.

**Figure 1.**
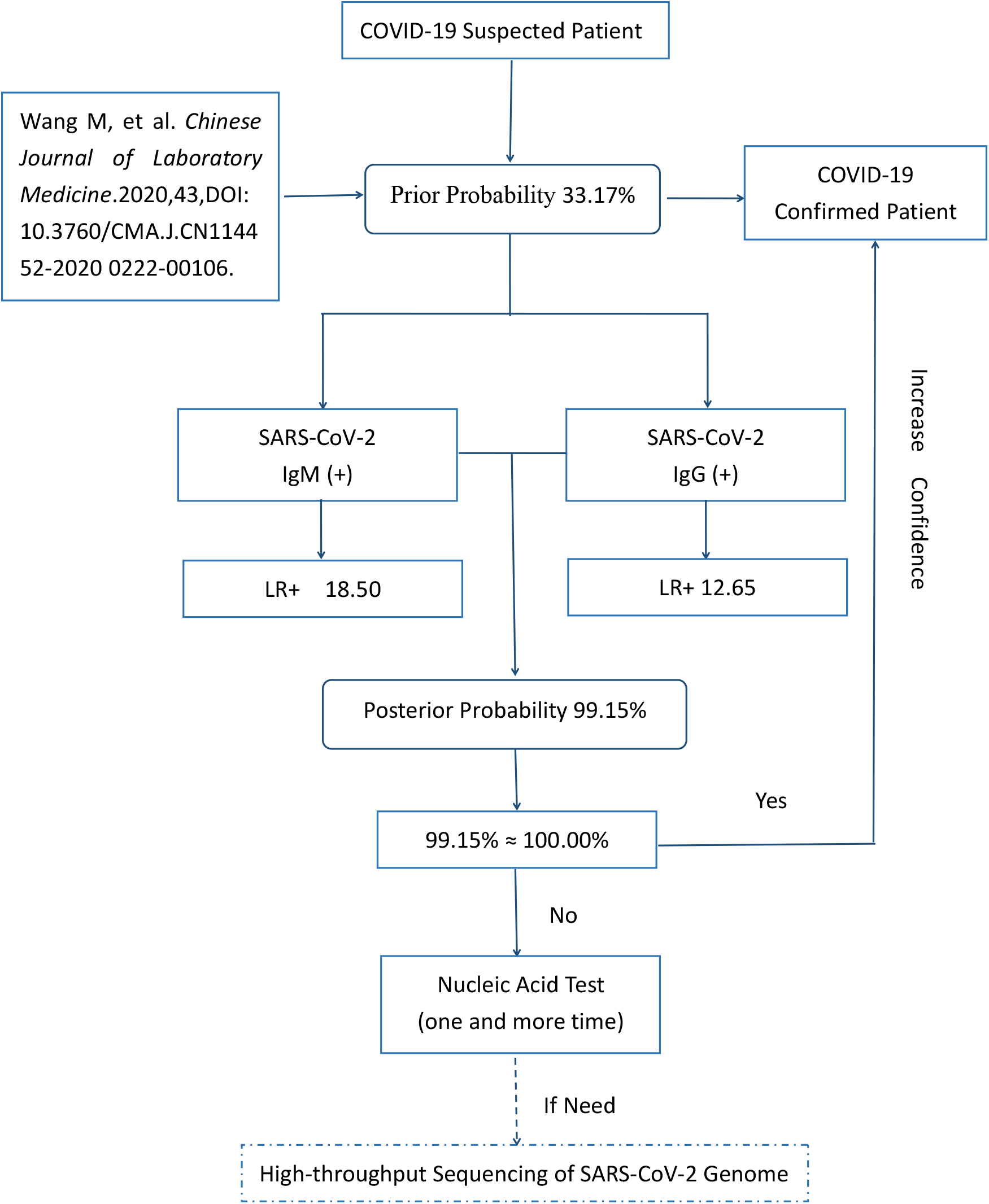
Clinical Pathway for Clinicians to Diagnose COVID-19.

## Discussion

With the continuous development of clinical laboratory’s detection level of antibody from human body after SARS-CoV-2 infection, in the “Diagnosis and Treatment Protocol for Novel Coronavirus Pneumonia (Trial Version 7)”, serological antibody detection is given the same status as nucleic acid detection for the first time when the diagnosis of COVID-19. Meanwhile, National Medical Products Administration of China has also approved 8 antibody detection kits in an emergency, laying the foundation for antibody detection to enter clinical practice.

Nucleic acid detection and antibody detection have their own advantages and disadvantages ^[6-7]^. First of all, from the aspect of specimen sampling, commonly used specimens for nucleic acid detection are sputum, nasopharynx swab or various lung lavage fluids. Sampling has great infection risks for medical personnel and also has potential infection risks for laboratory personnel for nucleic acid detection^[8]^. However, the type of antibody test specimen is blood, which is relatively easy to obtain and the infection risk of medical personnel is relatively low. Secondly, according to the quality of detection results, nucleic acid detection is restricted by the quality of sample materials and the extraction quality of nucleic acid, and there is a serious undetected phenomenon. Different sample types have different detection rates, Sucha as bronchoalveolar lavage fluid specimens showed the highest positive rates (93%), sputum (72%), nasal swabs (63%), fibrobronchoscope brush biopsy (46%), pharyngeal swabs (32%), feces (29%), and blood (1%)^[9]^. So continuous nucleic acid detection for several times for some suspected patients is very necessary and can effectively avoid false negative result. For antibody detection, except for some immunocompromised patients, which cannot produce antibodies effectively, antibodies should be present in most infected persons^[10]^. Antibody detection is mainly limited by the sensitivity and related performance of the kit. In addition, there will be some false positives caused by detection interference in tumor patients, patients with autoimmune diseases and other patients with chronic infection. Finally, the convenience of detection operation, antibody detection has advantages that nucleic acid detection does not have at all, including simplicity and convenience of operation, qualification of personnel, requirements of detection environment, etc. Therefore, it has great clinical value to study how to optimize antibody detection and nucleic acid detection in clinical diagnosis.

Through searching the relevant articles, no clear research about the optimization path of antibody and nucleic acid detection is found. Therefore, this research uses the published data and adopts the method of positive likelihood ratio to calculate the posterior probability to propose the possibility of optimizing the diagnosis path for the first time.Since the original result data of the two antibodies tandem test cannot be obtained from “Diagnostic Value of Combined Detection of Serum 2019 novel coronavirus IgM and IgG Antibodies in novel coronavirus Infection”, the diagnostic performance index of tandem test cannot be analyzed in this paper. However, this study solves the problem by calculating the posterior probability through the LR+, and proves that the posterior probability of the two antibodies tandem test is much higher than each single antibody detection, while the posterior probability of single antibody detection is not much different. This shows that the single antibody test does not significantly increase the confidence of clinicians in the clear diagnosis of confirmed cases from suspected cases. However, the two antibodies tandem test obviously improved this confidence, making the clinician’s confidence in diagnosis increased to 99.15%,which is helpful for clinicians to optimize the diagnosis process by quantification method. By the study, the clinical pathway for clinicians to diagnose COVID-19 was that, for suspected cases, IgM and IgG antibody tests should be firstly done at the same time. If the antibody tests are all positive, COVID-19 can be confirmed. If not, nucleic acid detection (one or more times) is performed, and in extreme cases, high-throughput viral genome sequencing is performed^[11]^.

In short, using positive likelihood ratio to calculate the posterior probability could better reflect the tandem detection of the two antibodies compared with conventional diagnostic performance indicators. In practical work, it is more helpful for clinicians to optimize the diagnosis process. In addition, this study is aimed at the secondary excavation of published literature data. In the future, the original data should be used and the sample size expanded to further verify the conclusion.

## Data Availability

all data are publicly available，Which can contact the corresponding author for the article data.

